# Low zinc levels at clinical admission associates with poor outcomes in COVID-19

**DOI:** 10.1101/2020.10.07.20208645

**Authors:** Marina Vogel-González, Marc Talló-Parra, Víctor Herrera-Fernández, Gemma Pérez-Vilaró, Miguel Chillón, Xavier Nogués, Silvia Gómez-Zorrilla, Inmaculada López-Montesinos, Judit Villar, Maria Luisa Sorli-Redó, Juan Pablo Horcajada, Natalia García-Giralt, Julio Pascual, Juana Díez, Rubén Vicente, Robert Güerri-Fernández

**Affiliations:** Laboratory of Molecular Physiology, Department of Experimental and Health Sciences, Universitat Pompeu Fabra, Barcelona, Spain; Molecular Virology Group, Department of Experimental and Health Sciences, Universitat Pompeu Fabra, Barcelona, Spain; Department of Biochemistry and Molecular Biology and Institute of Neurosciences, Edifici H, Universitat Autònoma de Barcelona, E-08193, Bellaterra, Spain; Unitat Mixta UAB-VHIR, Vall d’Hebron Institut de Recerca (VHIR), Barcelona, Spain; Institut Català de Recerca i Estudis Avançats (ICREA), Barcelona, Spain; Institut Mar d’Investigacions Mediques, Barcelona, Spain; Department of Internal Medicine, Hospital del Mar, Autonomous University of Barcelona, Spain; CIBERFES, Instituto Carlos III, Madrid, Spain; Institut Mar d’Investigacions Mediques, Barcelona, Spain; Department of Infectious Diseases, Hospital del Mar, Autonomous University of Barcelona, Spain; CIBERFES, Instituto Carlos III, Madrid, Spain; Institut Mar d’Investigacions Mediques, Barcelona, Spain; Department of Nephrology, Hospital del Mar, Autonomous University of Barcelona, Spain

## Abstract

**Background:** Biomarkers to predict Coronavirus disease-19 (COVID-19) outcome early at infection are urgently needed to improve prognosis and treatment. Zinc balances immune responses and also has a proven direct antiviral action against some viruses. Importantly, zinc deficiency (ZD) is a common condition in elderly and individuals with chronic diseases, two groups with more severe COVID-19 outcomes. We hypothesize that serum zinc content (SZC) influences COVID-19 disease progression and thus might represent a useful biomarker.

**Methods:** We run a retrospective observational study with 249 COVID-19 patients admitted in Hospital del Mar. We have studied COVID-19 severity and progression attending to SZC at admission. In parallel we have studied SARS-CoV2 replication in the Vero E6 cell line modifying zinc concentrations.

**Findings:** Our study demonstrates a correlation between serum zinc levels and COVID-19 outcome. Serum zinc levels lower than 50 µg/dl at admission correlated with worse clinical presentation, longer time to reach stability and higher mortality. Our *in vitro* results indicate that low zinc levels favor viral expansion in SARS-CoV2 infected cells.

**Interpretation:** SZC is a novel biomarker to predict COVID-19 outcome. We encourage performing randomized clinical trials to study zinc supplementation as potential prophylaxis and treatment with people at risk of zinc deficiency.

**Funding:** Spanish Ministry of Science and Innovation, “Maria de Maeztu” Programme for Units of Excellence in R&D and Secretaria d’Universitats i Recerca del Departament d’Economia i Coneixement of the Generalitat de Catalunya. Instituto Carlos III Fondos de Investigaciones Sanitarias (FIS), CIBER on Frailty and Healthy Ageing and FEDER funds

## INTRODUCTION

Infections with SARS-CoV-2 result in a systemic disease with a variety of outcomes, from no symptoms to severe and diverse pathologies, being pneumonia and acute distress the most common. Therefore, biomarkers to predict disease severity in early infection stages are urgently needed.

Zinc (Zn^2+^) is an essential trace element required for maintaining a variety of fundamental biological processes due to its functions as a cofactor, signaling molecule, and structural element. One of the most significant roles of zinc in our body is its broad effect on the immune system^1,2^, as both, the adaptive and the innate immunity, are affected by zinc levels. Consequently, zinc deficiency (ZD), due to low zinc intake and/or malabsorption, results in an immune imbalance that ultimately causes a major public health problem with high prevalence in elderly and individuals with chronic diseases^1^. In adaptive immunity, zinc affects T lymphocyte maturation, differentiation and cytokine production. B cells activation and plasma cell differentiation depend on zinc signaling as well^2,3^. In innate immunity, zinc has an anti-inflammatory role^4^. Concretely, ZD is associated with higher levels of IL-1beta and TNF-alfa (5) and with altered activities of monocytes, neutrophils and NK cells^4,6,7^. Correspondingly, ZD results in increased susceptibility to inflammatory and infectious diseases including acquired immune deficiency syndrome, measles, malaria, tuberculosis, and pneumonia (Reviewed in^2^).

In this context, zinc supplementation trials have been carried out to reduce morbidity and mortality in developing countries with high ZD incidence. Importantly, zinc supplementation significantly reduced the duration of respiratory tract infections caused by rhinoviruses and coronaviruses^8^. Moreover, zinc has been shown to have a direct antiviral action (reviewed in ^9^). Remarkably, zinc inhibited *in vitro* both, the binding of the SARS coronavirus RNA-dependent RNA polymerase to its template and the subsequent elongation, and viral replication in cell culture^10^. Altogether, zinc status might condition COVID-19 severity and zinc supplementation could be a useful tool to impact COVID-19 outcome.

In this work we focus our observational study on the putative association between zinc status of hospitalized COVID-19 patients with disease progression and clinical outcomes. Moreover, we address in cell culture the potential of zinc supplementation to directly block SARS-CoV-2 multiplication in Vero E6 infected cells.

## METHODS

### Study design and participants

A retrospective observational cohort study was performed at Hospital del Mar in Barcelona (Spain). This is a 400-bed university tertiary hospital in Barcelona that provides healthcare to an urban area of 500,000 people. All patients admitted with COVID-19 for ≥ 48h between 9^th^ March and 1^st^ April 2020 were included. COVID-19 was defined as a SARS-CoV-2 infection confirmed by quantitative PCR (qPCR) performed in nasopharyngeal samples obtained by trained personnel at hospital admission, and/or by fulfilling clinical diagnostic criteria. These include any of the following: respiratory symptoms (dyspnea, cough, sore throat, changes in taste/smell) chest X-Ray findings (uni- or bilateral interstitial infiltrates) that made the diagnosis probable in the current epidemiological situation. The Institutional Ethics Committee of Hospital del Mar of Barcelona approved the study and due to the nature of the retrospective data review, and waived the need for informed consent from individual patients (CEIm 2020/9352).

### Procedures

Demographic, clinical, epidemiological and the whole episode (laboratory workout, vital signs, treatment) data were extracted from the electronic medical record using a standardized data collection method.

Laboratory workouts were systematized with an at-admission protocol that included a fasting blood draw complete kidney and liver profile, electrolytes, blood count, coagulation profile, inflammatory markers (interleukin-6 (IL-6), serum ferritin), D-dimer, myocardial enzymes, zinc and selenium levels.

### Definitions

#### Time to clinical stability (TCS)

TCS was defined as the time elapsed since the patient’s admission to: oxygen saturation >94% (FiO2 21%), normalized level of consciousness (baseline), Heart Rate <100rpm, systolic Blood Pressure > 90mm Hg, temperature <37·2C.

#### Clinical severity

Clinical severity was assessed at admission with Modified Early Warning Score (MEWS)^11^. The same score was used for the follow-up during the admission.

#### Zinc deficit

According to previous publications the threshold of Zn^2+^ considered abnormally lower was 50 µg/dl (7·6 µM) (12) This cutoff was set to categorize Zn^2+^ levels. We considered as predictive factor the lower range (<50 µg/dl).

### Cell culture

Vero E6 cells were grown as described^13,14^. When indicated, FBS was incubated according to the manufacturer’s instructions with Chelex 100 resin (Bio-Rad Laboratories) to generate Zn^2+^-free growth medium. ZnSO_4_ was added as needed to the final medium to generate specific Zn^2+^ concentration conditions. Chloroquine (Supelco, #PHR1258) was prepared in water at 10 mM and used at the desired concentration.

### Zinc measurements

Cells were seeded and grown in multi well 24 plates until reaching 80% of confluence. Cells were incubated with 1 µM of FluoZin-3AM (Invitrogen) or 25 µM of Zinquin (Sigma-Aldrich) for 30 min at 37°C (5% CO_2_) in isotonic solution containing (in mM) 140 NaCl, 5 KCl, 1.2 CaCl_2_, 0·5 MgCl_2_, 5 glucose, and 10 Hepes (300 milliosmoles/liter, pH 7·4) plus different concentrations of Zn^2+^ and/or Chloroquine (CQ). Cells were then dissociated with Trypsin 0·05% in 0·53 mM EDTA and were washed with PBS 1x. Fluorescence was quantified using a LSRII flow cytometer. Further analysis was performed using Flowing Software (Perttu Terho).

For in vivo confocal imaging, cells growth on 22mm coverslips were incubated with Lysotracker together with FluoZin-3AM or Zinquin in a solution with 50 µM Zn^2+^ for 30min, washed twice with PBS and placed under the microscope in isotonic solution for imaging with a SP8 Leica microscope.

### Viability assays

Cells were exposed to different Zn^2+^ and CQ concentrations for 48h. Then, MTT reagent was added to obtain a final concentration of 0·5 mg/ml. Cells were incubated 2-3 h at 37°C. After that, supernatant was removed and cells were resuspended in 100 µl of DMSO. The absorbance was read at 570 nm.

### Virus infection and quantification

Vero E6 cells were infected with SARS-CoV-2 strain hCoV-19/Spain/VH000001133/2020 (EPI_ISL_418860) and 48 h later viral RNA from the supernatant was extracted using the Quick-RNA Viral Kit (Zymo Research). SARS-CoV-2 production was quantified by qPCR with the qScript™ XLT One-Step RT-qPCR ToughMix®, ROX™ (Quanta Biosciences) using the following specific probe: 2019-nCoV_N1-P, 5’-FAM-ACCCCGCATTACGTTTGGTGGACC-BHQ1-3’; and primers: 2019-nCoV_N1-F, 5’-GACCCCAAAATCAGCGAAAT-3’; and 2019-nCoV_N1-R, 5’-TCTGGTTACTGCCAGTTGAATCTG-3’ (Biomers).

### Western blot

Cells were treated with 0, 10 or 50 μM of Zn^2+^ and 10 μM of CQ for 24 h. Then, cells were washed twice with cold PBS and lysed with 35 μl of lysis buffer (50□M Tris-HCl pH 7·4, 150□M NaCl, 5□M EDTA, 0.5% NP-40, 1□M DTT, 10□M 13-GP, 0·1□M Na_3_VO_4_ and protease inhibitors). Lysates were vortex for 30 min at 4°C and centrifuged at 10000 x *g* to remove aggregates. Lysates were boiled at 95 °C and placed 1 min in ice. 20 μl of each sample were loaded onto a 12 or 14 % polyacrylamide gel. After transfer, membranes were blocked with 5% of milk in TBS-Tween 0·1% for 1 h at room temperature. Primary antibodies were diluted in blocking solution: LC3 (L8918, Sigma) at 1:500; p62 (ab155686, Abcam) at 1:1000; GAPDH (ab8245, Abcam) at 1:1000; and incubated overnight at 4°C. Anti-rabbit or anti-mouse HRP secondary antibodies (1:1000; GE Healthcare) were used.

### Statistical analysis

A descriptive analysis of the main demographic characteristics of the cohort, and the main clinical (laboratory, treatment and outcome) characteristics of the episode was done. Continuous and categorical variables were presented as median (interquartile range (IQR)) and absolute number (percentage), respectively. Mann-Whitney U-test, χ2 test and Fisher’s exact test were used to compare differences between individuals with serum zinc levels above and below 50 µg/ml respectively. Bi-variate comparisons and a multiple logistic regression model studying the impact of at admission serum zinc in in-hospital mortality were fitted adjusting by age, sex, comorbidities and severity of the episode. Taking into consideration the number of deaths observed and to avoid overfitting the model we selected the most significant variables and those that provided a more general characterization of the individuals. We excluded variables if they had collinearity with MEWS, or age. Significance was a p-value <0.05. Stata 14 software was used.

In the *in vitro* assays, we applied either unpaired Student’s *t* test (when comparing two conditions) and one-way analysis of variance (ANOVA) followed by Bonferroni post hoc test (when comparing control with any other condition) were applied. All plots comparisons and p values are described in figure legends. Analysis was performed using GraphPad software.

## RESULTS

We included 249 consecutive adults admitted to the COVID-19 unit between March 9^th^ and April 1^st^, 2020. The median age of participants was 65 (54-75) years, 49% were female. Table 1 shows the baseline characteristics of the cohort and the differences between individuals with SZC < 50 µg/dl and *≥* 50 µg/dl at onset. Hypertension was the most prevalent comorbidity. At admission the overall severity according to MEWS score was 2 (1-3). Fever, cough and dyspnea were the most common symptoms with a frequency of 203 (81%), 198 (79%) and 151 (60%) respectively. Inflammation was a hallmark of the episode with 224 (87·5%) of individuals with an abnormal IL-6 at admission and with a median IL-6 42pg/ml (15-89), a median C-Reactive-Protein 7·5 mg/dl (3·5-15.2), and a median D-Dimer 800 UI/l (470-1450). Almost all the individuals admitted during this period received hydroxychloroquine. Other treatments were tocilizumab, dexamethasone and methylprednisolone that were prescribed in a quarter of individuals each one. 70 patients (28%) were admitted to the ICU and 21 (9%) patients died during hospitalization in this period.

SZC median at admission were 61 µg/dl (9·3 µM) with 58 (23%) of the individuals presenting SZC <50 µg/dl. Individuals with serum zinc levels <50 µg/dl had higher prevalence of chronic kidney disease and chronic respiratory disease 9% vs 2%; p=0·024 and 16% vs 7%; p=0·041. Moreover, this group had a more severe clinical presentation MEWS score (2 (2-3) vs 2 (1-3); p=0·005) and a significantly higher inflammation measured by the nonspecific marker C-Reactive protein 14·6mg/dl vs 7mg/dl; p=0·03 and the more specific one IL-6 77pg/ml vs 32pg/ml; p<0·001. At admission, SZC correlated negatively with inflammation measured by IL-6 (Pearson’s r= −0·307; p<0·001) (Fig 1A) and, interestingly also correlated with the highest value of IL-6 during the episode (Pearson’s r=-0·317;p<0·001) (Fig 1B). This negative correlation was also observed with the nonspecific inflammatory markers such as ferritin (Pearson’s r=-0·227; p=0·001) and C-Reactive Protein (Pearson’s r=-0·315; p<0·001) (Fig 1C). We also found correlation between SZC at admission and procoagulation factors such as D-dimer (Pearson’s r=-0·317; p=0·048).

**Figure 1.**
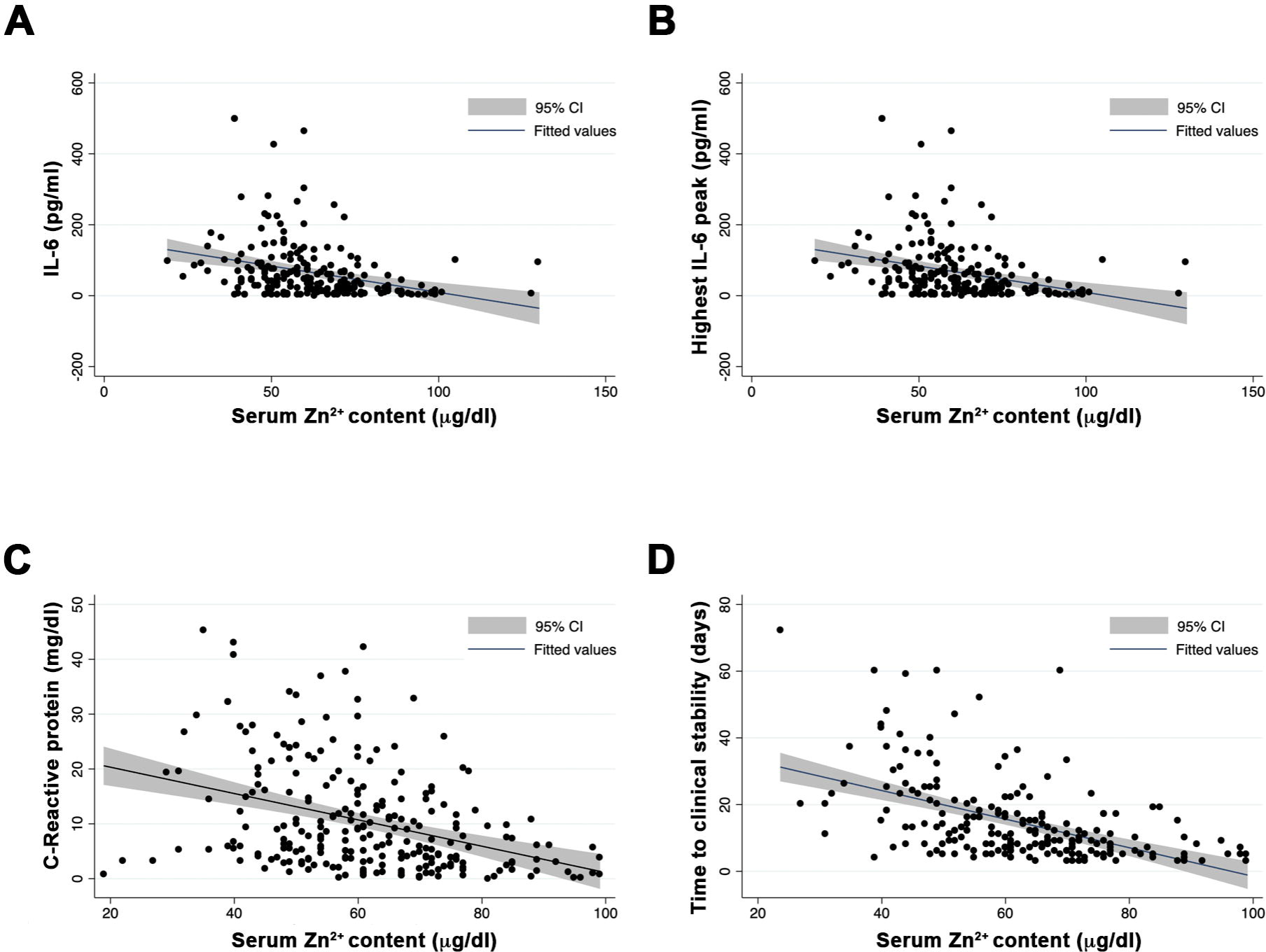
Correlation between serum zinc levels at admission, inflammatory markers and clinical outcome. A. Zinc and IL-6 at admission. B. Zinc at admission and highest value of IL-6 during the episode. C. Zinc and C-Reactive Protein at admission. D. Serum zinc content at admission and time to clinical stability.

The median time to reach clinical stability in our cohort was 9·5 days (6-18). However, subjects with SZC at admission <50 µg/dl needed longer TCS stability compared with those with zinc *≥*50 µg/dl, 25 days (14-36) vs 8 (5-14) days, respectively; p<0·001. Moreover, SZC at admission was correlated negatively with TCS of the episode (Pearson’s r=-0·441;p<0·001) (Fig 1D). In a multivariable linear regression model adjusted by age, sex, severity and comorbidities TCS was associated with low SZC at admission (beta-coefficient −0·21 (95% CI −0·316 to −0·097; p<0·001)). In an alternative model, serum zinc levels <50 µg/dl negatively impacted on TCS (beta-coefficient 14·1 (95% CI 4·29 - 23·94; p=0·004).

Twenty-one individuals (9%) died during this period. SZC at admission was significantly higher among individuals who survived 62 µg/dl (52-72) compared to those who died 49 µg/dl (42-53); p<0·001. Individuals with SZC at admission <50 µg/dl had a mortality of 21% that was significantly higher compared with 5% mortality in individuals with zinc at admission ≥50 µg/dl; p<0·001. We fitted a multivariable logistic regression model including 249 patients with data for all variables (228 survivors and 21 non-survivors). When adjusting by age, sex, Charlson comorbidity index and severity, the model showed an odds ratio (OR) for in-hospital death of 0·94 [95% CI 0·899 to 0·982; p=0·006) per unit increase of serum zinc at admission. In an alternative adjusted model with the predictive variable SZC at admission <50 µg/dl the adjusted OR for *in-hospital* death was 3·2 (95% CI 1·01 to 10·12; p=0·047).

To further understand the link between zinc status and COVID-19 severity, we carried out assays in cell culture to study the direct impact of cellular zinc content on SARS-CoV-2 multiplication. We used three different concentrations of zinc in the extracellular medium, 0, 10 and 50µM ZnSO_4_, to reproduce zinc deficiency, physiological zinc and zinc supplementation, respectively. As expected, intracellular Zn^2+^ content changed in Vero E6 cells incubated for 30min in different extracellular Zn^2+^ concentrations and monitored with Fluozin 3AM and Zinquin fluorescence probes. Confocal images showed that FluoZin 3AM signal was mainly localized in the lysosomal compartment while Zinquin signal was intracellularly spread (Fig 2B). After 48h incubation, different Zn^2+^ content solutions did not impact cell viability (Fig 2C). Importantly, extracellular zinc concentrations affected SARS-CoV-2 multiplication as measured by qPCR from the cell culture supernatant at 48h post infection. SARS-CoV-2 multiplication was increased at 0 µM Zn^2+^ when compared to those values at 10 and 50 µM Zn^2+^ concentrations (Fig 2D). This indicates that zinc levels affect the SARS-CoV-2 life cycle in infected cells.

**Figure 2.**
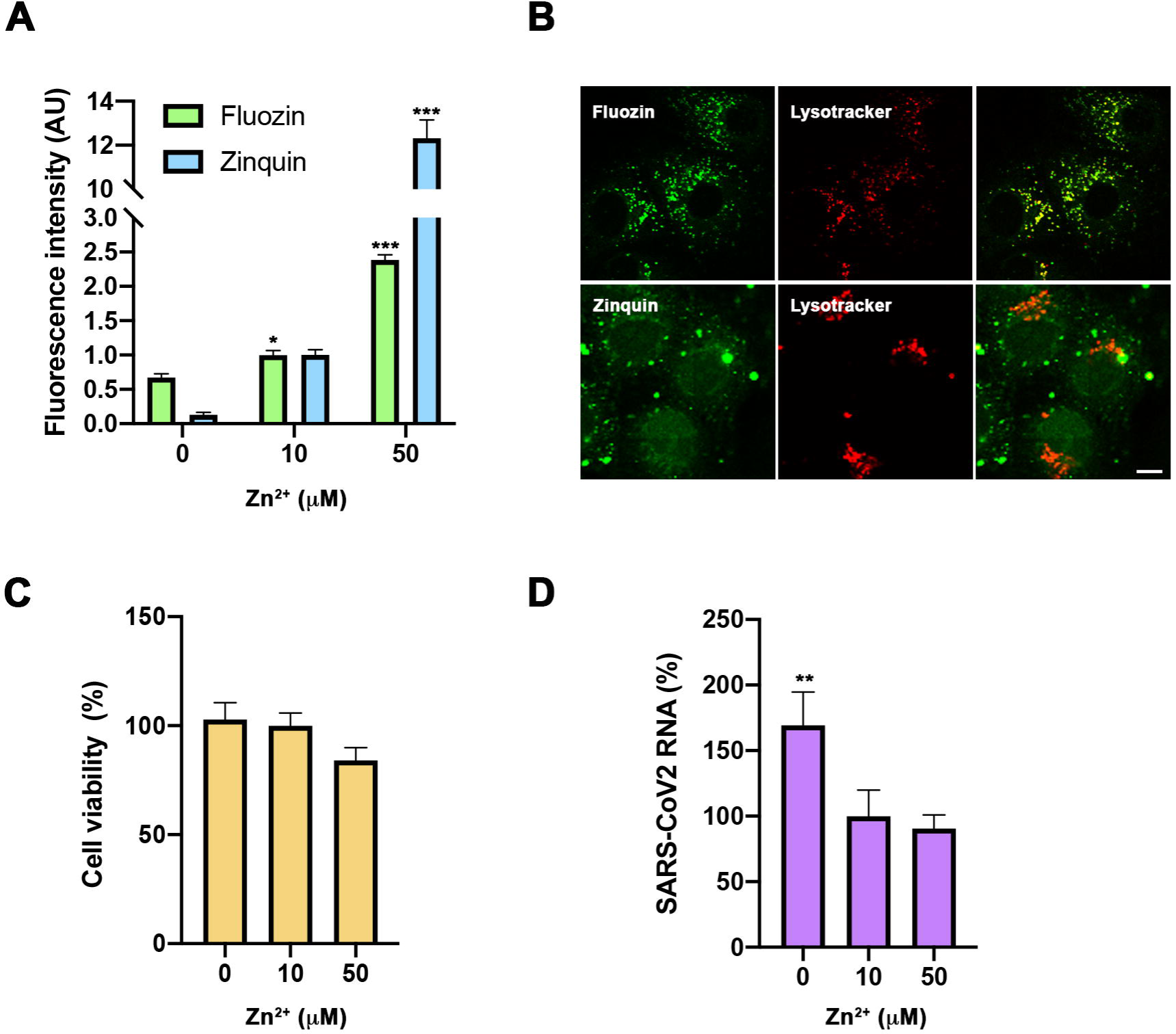
Evaluation of zinc homeostasis in SARS-CoV2 infection. A-B. Intracellular zinc content measurement using FluoZin-3AM and Zinquin probes in Vero E6 cells. A. Flow cytometry in cells incubated for 30min with 0, 10 and 50µM extracellular Zn^2+^ content. Intensity expressed in Arbitrary Units (AU) (n=3; * p<0·05, *** p<0·001; Bonferroni-corrected one-way ANOVA compared to 0 Zn^2+^). B. Confocal images of living cells incubated with FluoZin-3AM (up) or Zinquin (bottom) and Lysotyracker in 50µM Zn^2+^ extracellular medium. Scale bar 10µm. C. Viability MTT assay in cells incubated with 0, 10 and 50µM extracellular Zn^2+^ content for 48h. Data expressed in percentage compared to control condition (10µM Zn^2+^) (n=12; Bonferroni-corrected one-way ANOVA compared to 10µM Zn^2+^). D. Quantification in supernatant of viral RNA copies by qPCR in cells infected with SARS-CoV2 and collected at 48h. Data expressed in percentage compared to control condition (10µM Zn^2+^) (n=3; **p<0·01; Bonferroni ANOVA compared to 10µM Zn^2+^).

It has been suggested that Zn^2+^ may potentiate CQ antiviral activity^15^. To test this, we carried out SARS-CoV-2 infections at different zinc concentrations in the absence or presence of 10 µM CQ, the concentration chosen based on previous published effective concentration ^13,14^ Neither the CQ toxicity nor its antiviral activity were affected by zinc levels (Fig 3A,B). CQ caused a significant reduction in SARS-CoV-2 RNA genome copies compared to control cells (59·68±18·46%; p<0.05) (Fig 3B). Next, we addressed whether CQ increases the intracellular Zn^2+^ content by acting as a zinc ionophore, as was previously proposed^16^. For this, we evaluated the cytosolic Zn^2+^ levels in cells growth under different CQ concentrations using flow cytometry analysis and FluoZin-3AM and Zinquin labels (Fig 3C). Notably, an increased zinc signal was observed in a CQ dose dependent manner using FluoZin-3AM but not with Zinquin (Fig 3C). This indicates that CQ modifies lysosomal Zn^2+^ content but it is not a zinc ionophore. As the described increase in lysosomal pH caused by CQ blocks autophagic flux (17) and inhibition of autophagy impairs SARS-CoV-2 replication ^18^, we studied the impact on autophagy of different zinc concentrations in the presence and the absence of CQ (Fig 3D-F). As expected, at 10µM CQ treatment autophagy blockade resulted in an increased LC3II/I ratio and p62 expression (Fig 3E,F). However, no significant effects on these values were observed at different Zn^2+^ levels in the absence or presence of CQ. Altogether, our results indicate that CQ and zinc do not potentiate each other.

**Figure 3.**
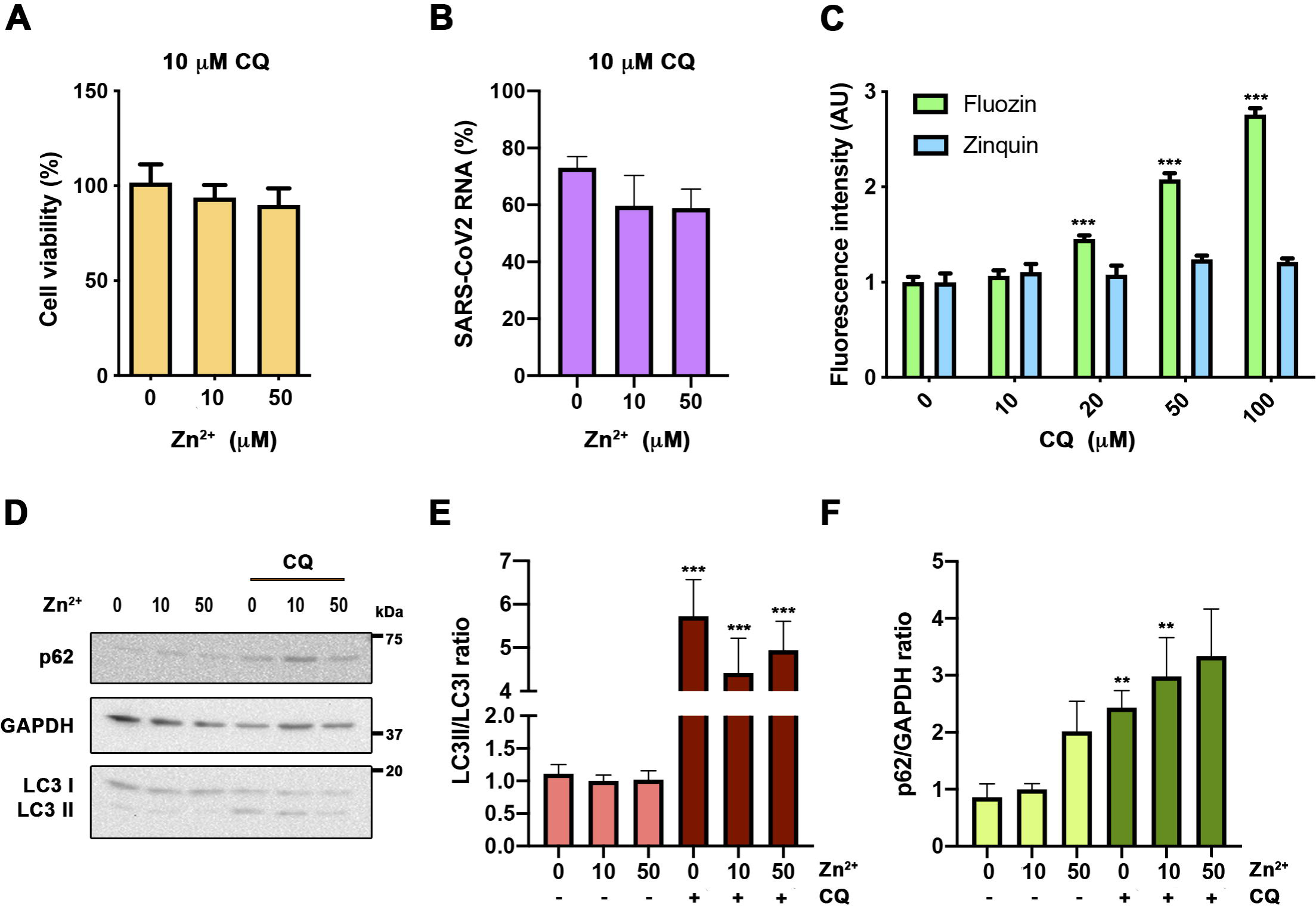
Evaluation of zinc potentiation of chloroquine antiviral action. A. Viability MTT assay in cells incubated with 10µM CQ in 0, 10 and 50µM Zn^2+^ content for 48h (n=15; Bonferroni-corrected one-way ANOVA compared to 10µM Zn^2+^). B. Quantification in supernatant of viral RNA copies by qPCR in cells infected with SARS-CoV2 treated with 10µM CQ in 0, 10 and 50µM Zn^2+^ and collected at 48h. Data expressed in percentage compared to control condition (10µM Zn^2+^ without CQ, Fig 2D) (n=3; Bonferroni-corrected one-way ANOVA compared to 10µM CQ in 10µM Zn^2+^). C. Flow cytometry in cells incubated for 30min with 0, 10 and 50µM extracellular Zn^2+^ content using FluoZin-3AM and Zinquin probes. Intensity expressed in Arbitrary Units (AU) (n≥4; *** p<0·001; Bonferroni-corrected one-way ANOVA compared to 0µM Zn^2+^ from each probe). D-F. Western blot analysis from cells incubated for 24h in 0, 10 and 50µM Zn^2+^ with or without 10µM CQ. Antibodies against LC3, p62 and GAPDH. D. Representative blot. E-F. Quantification of the bands for LC3 (E; n≥10) and p62 (F; n≥7). (Bonferroni-corrected one-way ANOVA compared to 10µM Zn^2+^ or 10µM CQ in 10µM Zn^2+^. ** p<0·01, *** p<0·001 unpaired t-test comparing conditions with and without CQ).

## DISCUSSION

Our study demonstrates a correlation between serum zinc levels and COVID-19 outcome. SZC lower than <50 µg/dl at admission correlated with worse clinical presentation, longer time to reach stability and higher mortality. These results point at SZC as an early predictor of COVID-19 severity, whose adjustment might also constitute an early therapeutic intervention point. Moreover, we show that SZC affects the SARS-CoV-2 life cycle in the infected cells. This effect, contrary to what it was previously suggested, does not seem to potentiate CQ activity.

The association between SZC and human health is known for decades^1^. Due to poor nutrition and subsequent low zinc intake, ZD remains a major nutritional problem in multiple countries. In addition, elderly individuals are prone to ZD even in developed countries where the incidence ranges from 15 to 31% depending on the age and the country of study^19^. Older adults are the group at higher risk for severe symptoms and mortality from COVID-19. In our retrospective observational study with 249 COVID-19 patients admitted to Hospital del Mar, 23% of them had at admission SZC lower than 50µg/dl, the cutoff associated with ZD and development of clinical signs^12^.

At onset, higher levels of inflammatory markers such as IL-6 and C-reactive protein were present in low SZC patients (Fig 1A,C). The prognostic value for COVID-19 severity of IL-6 and C-reactive protein has already been described^13^. In this work we highlight the importance of SZC as an early predictor of COVID-19 severity and clinical outcome, and consequently, as a potential therapeutic intervention point. ZD is known to be associated with proinflammatory responses at infection, showing higher reactive oxygen species production and inflammatory markers^20,21^. An imbalance in cytokine production by cells of both innate and adaptive immunity has also been reported^3,2^. In our study, we observed a robust negative correlation between zinc levels and IL-6 (Fig 1B). In this respect, it has been reported that IL-6 potentiates SZC decrease by promoting zinc internalization in hepatocytes in acute phase of infection^22^. We cannot discard that the hypozincemia observed in COVID-19 patients might be caused and worsened by a negative IL-6 production loop.

Several clinical trials using zinc supplementation have been carried out to prevent and treat infections and inflammatory conditions^1,2^. Zinc supplementation decreases the incidence of infection in elderly and improves cytokine imbalance and oxidative stress markers^23,24^. Thus, the idea of supplementing low SZC COVID-19 patients with zinc in order to balance what has been called the cytokine storm caused by SARS-CoV-2 is attractive. Reassuringly, zinc supplementation for the common cold caused by rhino- and coronaviruses has been proven to reduce its duration and symptoms^8^. In addition to the impact of zinc in the modulation of the antiviral immune response, zinc has also been shown to have a direct antiviral action^9^. We have analyzed *in vitro* the impact of zinc homeostasis in SARS-CoV-2 infection. Our results indicate that hypozincemia favors viral expansion in the infected cell (Fig 2C). These results would support that the poor clinical outcome observed in low SZC patients is caused by the effect of ZD on both, inducing immune imbalance and increasing viral load via promoting viral expansion in the infected cell. Of note, our *in vitro* studies, however, did not show a replication blockade in zinc supplementation conditions (50µM, Fig 2C) suggesting the need of a zinc ionophore to further increase cytosolic zinc levels as previously shown in vitro for Herpes simplex, picornavirus, arterivirus and SARS-CoV coronavirus^9,10^.

During the beginning of the pandemic CQ was prescribed as a first line treatment in the acute presentation. The rationale was that CQ has been claimed to be a novel zinc ionophore^16^ and CQ showed antiviral effects against SARS-CoV-2 *in vitro*^13,14^. However, clinical trials failed to demonstrate beneficial effect over COVID-19^25,26^. Therefore, it was suggested that zinc supplementation could potentiate its antiviral activity^15^. In our results *in vitro*, cellular zinc content did not modify CQ cytotoxicity, the autophagic flux blockade or its antiviral action against SARS-CoV2. Moreover, we monitored intracellular zinc content both with Fluozin-3AM, a probe previously done by Xue and colleagues that is retained mainly in lysosomes^16^, and with Zinquin, which presents a more general intracellular staining. We conclude that CQ is not a zinc ionophore as previously claimed because its effect on cellular zinc content is restricted to the lysosomal compartment, probably by altering zinc transport at this specific organelle (Fig 3). Thus, our study does not support the mechanistic rationale of supplementing CQ treatments with zinc.

This work aims to focus clinical attention on serum zinc content in COVID-19 patients. Our analysis has shown a robust correlation between low SZC and COVID-19 severity and mortality. The cause is likely to be a combination of immune system imbalance and a direct benefit of viral replication. Thus, we propose SZC as a novel parameter to predict COVID-19 outcome. It is then urgent to start clinical trials supplementing with zinc low SZC patients at admission to reestablish zinc homeostasis. It should be also recommended to promote zinc supplementation programs targeted to people at risk of zinc deficiency, such as elderly, in order to reduce COVID-19 severity.

## Data Availability

data will be available once this study has been published.

## ACKNOWLEDGEMENTS

This work was supported by the Spanish Ministry of Science and Innovation through grants PID2019-106755RB-I00/AEI/10.13039/501100011033 to RV and PID2019-106959RB-I00/AEI /10.13039/501100011033 to JD, an institutional “Maria de Maeztu” Programme for Units of Excellence in R&D (CEX2018-000792-M) to RV and JD and by the 2017 SGR 909 grant from the Secretaria d’Universitats i Recerca del Departament d’Economia i Coneixement of the Generalitat de Catalunya to JD. RGF received support and funding from Centro de Investigación Biomédica en Red de Fragilidad y Envejecimiento Saludable (CIBERFES) [Grant number CB16/10/00245], FEDER funds and the FIS Project from Instituto de Salud Carlos III, Ministerio de Ciencia e Innovación [Grant number (PI19/00019)]

