## Supplementary material for "Low zinc levels at clinical admission associates with poor outcomes in COVID-19": Table 1

Table 1. Main clinical characteristics of the cohort. Differences between individuals with plasma zinc at admission <50mcg/dl and *≥50mcg/dl.*

|  | | *Overall* | | *<50* µg/*/dl* | |  | *≥50* µg/*/dl* | | *p-value* | |
| --- | --- | --- | --- | --- | --- | --- | --- | --- | --- | --- |
| **Cohort Characteristics** | | n=249 | |  | n=58 |  | n=191 | |  | |
| Median age, years (IQR) | | 65 | (54-75) * | 65 | (59-75) |  | 64 | (53-74) | | 0.363 |
| Male sex (%) | | 127 | (51%) | 30 | (51%) |  | 97 | (51%) | | 0.929 |
| Barthel index at admission, median (IQR) | | 100 | (100-100) | 100 | (100-100) |  | 100 | (100-100) | | 0.448 |
| Homelessness (%) | | 2 | (0.89%) | 0 | (0%) |  | 2 | (1.53%) | | 0.231 |
| **Comorbidities** | |  |  |  |  |  |  |  | |  |
| Current smoker (%) | | 23 | (9.3%) | 4 | (7%) |  | 19 | (10%) | | 0.482 |
| Hypertension (%) | | 141 | (56%) | 33 | (57%) |  | 108 | (57%) | | 0.962 |
| Diabetes Mellitus (%) | | 53 | (21%) | 17 | (29%) |  | 36 | (19%) | | 0.088 |
| Chronic lung disease (%) | | 22 | (9%) | 9 | (16%) |  | 13 | (7%) | | 0.041 |
| Chronic heart disease (%) | | 37 | (14%) | 11 | (19%) |  | 26 | (14%) | | 0.315 |
| Chronic renal disease (%) | | 70 | (12%) | 22 | (38%) |  | 48 | (25%) | | 0.058 |
| Chronic liver disease (%) | | 18 | (7%) | 1 | (2%) |  | 17 | (9%) | | 0.065 |
| Dementia (%) | | 8 | (3%) | 2 | (3%) |  | 6 | (3%) | | 0.908 |
| HIV-infection (%) | | 5 | (2%) | 1 | (2%) |  | 4 | (2%) | | 0.865 |
| Active cancer (%) | | 7 | (3%) | 3 | (5%) |  | 4 | (2%) | | 0.214 |
| ACE inhibitors (%) | | 61 | (24%) | 12 | (21%) |  | 49 | (26%) | | 0.441 |
| Charlson Comorbidy Index, median (IQR) | | 1 | (0-3) | 1 | (0-3) |  | 1 | (0-2) | | 0.247 |
| **Symptoms at onset** | |  |  |  |  |  |  |  | |  |
| Dyspnoea (%) | | 151 | (60%) | 42 | (72%) |  | 109 | (57%) | | 0.036 |
| Fever (%) | | 203 | (81%) | 45 | (77%) |  | 158 | (82%) | | 0.377 |
| Cough (%) | | 198 | (79%) | 43 | (74%) |  | 155 | (81%) | | 0.246 |
| Diarrhea (%) | | 68 | (27%) | 12 | (21%) |  | 56 | (29%) | | 0.196 |
| Headache (%) | | 43 | (17%) | 8 | (14%) |  | 35 | (18%) | | 0.424 |
| Anosmia (%) | | 35 | (14%) | 7 | (12%) |  | 28 | (15%) | | 0.619 |
| **Clinical markers at onset** | |  |  |  |  |  |  |  | |  |
| Median C-Reactive Protein mg/dl (IQR) | | 7.5 | (3.5-15.2) | 14.6 | (5-24) |  | 7 | (3-13) | | 0.037 |
| Median lymphocyte count /ml (IQRS) | | 1.02 | (0.71-1.4) | 0.82 | (0.57-1.18) |  | 1.1 | (0.78-1.48) | | 0.598 |
| Median IL-6 pg/ml (IQR) | | 42 | (15-89) | 77 | (39-145) |  | 32 | (11-71) | | <0.001 |
| Median Lactate Dehydrogenase UI/l (IQR) | | 288 | (241-345) | 356 | (275-483) |  | 274 | (231-362) | | 0.001 |
| Median D-Dimer UI/l (IQR) | | 800 | (470-1450) | 935 | (540-1700) |  | 800 | (460-1215) | | 0.048 |
| Median PaFi (IQR) | | 177 | (100-299) | 124 | (91-181) |  | 219 | (106-314) | | <0.001 |
| Median MEWS (IQR) | | 2 | (1-3) | 2 | (2-3) |  | 2 | (1-3) | | 0.005 |
| Median serum Zinc, mcg/ml (IQR) | | 61 | (50-71) | 43 | (39-48) |  | 66 | (58-74) | | <0.001 |
| **Treatment** | |  |  |  |  |  |  |  | |  |
| Hydroxycholoroquine (%) | | 248 | (99.5%) | 58 | (100%) |  | 190 | (99%) | | 0.372 |
| Azythromicin (%) | | 231 | (95%) | 57 | (95%) |  | 174 | (95%) | | 0.766 |
| Tocilizumab (%) | | 55 | (23%) | 23 | (40%) |  | 32 | (17%) | | <0.001 |
| Dexamethasone (%) | | 64 | (26%) | 24 | (41%) |  | 40 | (21%) | | <0.001 |
| Methylprednisolone (%) | | 59 | (24%) | 26 | (45%) |  | 33 | (17%) | | <0.001 |
| **Clinical Outcomes** | |  |  |  |  |  |  |  | |  |
| Median Time to clinical stability days (IQR) | | 10 | (6-18) | 25 | (14-36) |  | 8 | (5-14) | | <0.001 |
| ICU admission (%) | | 70 | (28%) | 36 | (62%) |  | 34 | (18%) | | <0.001 |
| Death (%) | | 21 | (9%) | 12 | (21%) |  | 9 | (5%) | | <0.001 |
